# A Large Harmonized Upper and Lower Limb Accelerometry Dataset: A Resource for Rehabilitation Scientists

**DOI:** 10.1101/2024.08.15.24312066

**Authors:** Allison E. Miller, Keith R. Lohse, Marghuretta D. Bland, Jeffrey D. Konrad, Catherine R. Hoyt, Eric J. Lenze, Catherine E. Lang

## Abstract

Wearable sensors can measure movement in daily life, an outcome that is salient to patients, and have been critical to accelerating progress in rehabilitation research and practice. However, collecting and processing sensor data is burdensome, leaving many scientists with limited access to such data. To address these challenges, we present a harmonized, wearable sensor dataset that combines 2,885 recording days of sensor data from the upper and lower limbs from eight studies. The dataset includes 790 individuals ages 0 – 90, nearly equal sex proportions (53% male, 47% female), and representation from a range of demographic backgrounds (69.4% White, 24.9% Black, 1.8% Asian) and clinical conditions (46% neurotypical, 31% stroke, 7% Parkinson’s disease, 6% orthopedic conditions, and others). The dataset is publicly available and accompanied by open source code and an app that allows for interaction with the data. This dataset will facilitate the use of sensor data to advance rehabilitation research and practice, improve the reproducibility and replicability of wearable sensor studies, and minimize costs and duplicated scientific efforts.

## Background & Summary

The sophistication of wearable sensor technology is rapidly evolving, including drastic improvements in measurement precision, ease of use, and ability to capture multiple physiologic signals.^1-3^ Coinciding with these advancements is the increase in number of studies and range of use cases for these technologies in rehabilitation research and other realms of medicine.^4^ Wearable sensors have expanded outcome assessment from traditional laboratory/clinical settings to patients’ real-world environments, allowing outcomes most salient to patients to be captured in an ecologically valid context.^5,6^ Wearable sensor technology has been critical to a number of important scientific findings in rehabilitation research. For example, application of these technologies identified a critical practice gap that patient improvements on in-clinic assessments may not translate to improvements in daily life.^5^ Change in steps per day, a sensor-derived variable, served as the primary outcome for a large randomized controlled trial providing critical new insights on how to improve physical activity in individuals with chronic stroke.^7^ Longitudinal protocols have shed new light on how these technologies can be leveraged to predict onset and monitor disability progression in degenerative conditions.^8-10^ Thus, the profound impact that wearable technologies have had – and will continue to have – on rehabilitation research is clear.^11,12^

Despite these advancements, wearable sensor technology remains primarily confined to specialized research laboratories due to two key barriers. The first includes the resources required to deploy, manage, and inspect data from these technologies.^6^ Financial resources are required to purchase sensors, associated software, and accessories. Trained personnel add to these costs and are needed to deploy, monitor, process, and perform inspections of data derived from wearable sensors. Adding to these challenges is the often-lengthy and computationally complex pipelines needed to process the large quantities of data recorded by wearable sensors.^13,14^ Conquering these barriers can exceed the bandwidth of research laboratories unfamiliar with the technology, leaving important research questions related to the use of wearable sensors unaddressed and decelerating progress in both rehabilitation research and clinical practice.

The second large barrier is the lack of available data to inform wearable sensor data collection protocols and processing procedures. When designing a study that uses wearable sensor technology as a data collection tool, researchers are often faced with questions, such as ‘how many recording days are needed?’, ‘how should the sensor variable(s) of interested be calculated?’, and, if deploying sensors in an atypical population, ‘how does the sensor variable(s) behave in a neurotypical cohort?’.^15,16^ These unknowns often result in researchers needing to collect pilot data to inform their data collection protocols or add a control arm to their study design. These additional steps can delay the onset of data collection, add significant costs and (often duplicated) effort, and ultimately contribute to the wide variability in wearable sensor data collection and processing protocols in rehabilitation research.

These barriers could be addressed by providing the field with a large dataset of wearable sensor data collected from individuals who represent a variety of conditions, levels of disability, and demographic backgrounds and would ideally include a wide range of sensor variables and recording days. To be most useful to the field, the dataset would need to be findable, accessible, interoperable, and reusable (FAIR) and include open source code to allow others to replicate its data processing procedures and adapt for their own purposes, if necessary.^17^ We have taken the first steps towards this goal by harmonizing wearable sensor data collected across eight study protocols over the past 10+ years. In all eight studies, participants wore bilateral wrist sensors, bilateral wrist and bilateral ankle sensors, or a unilateral ankle sensor. Sensors were worn in daily life (i.e., outside the clinic/lab), with the exception of one study in which participants were residing in a skilled nursing facility. The harmonized dataset includes 790 participants ages 0 – 90 years, with a variety of conditions, and 2,885 recording days. The dataset is accompanied by open-source code hosted on Zenodo^18^ as well as an R Shiny object to allow others to visualize and interact with the data (https://langlab.shinyapps.io/harmonized_data/). We anticipate this harmonized dataset could be helpful in a variety of scenarios, including to: (1) help inform the design of wearable sensor data collection and processing protocols, (2) understand how a specific sensor variable behaves across populations, (3) develop new research questions or confirm the scientific merit of a proposed research question, and (4) minimize costs and duplicated efforts by allowing the field to address wearable sensor-related research questions using a large, pre-existing dataset. The purpose of this article is to describe the harmonized dataset, including information about each of the individual studies and the procedures used to harmonize the data.

## Methods

Figure 1 provides a schematic for how the eight studies (i.e., data sources) were harmonized into a single dataset. The harmonization process involved five main steps: (1) identifying the data sources to be harmonized, (2) preparing the data for harmonization, (3) developing a harmonized vocabulary, (4) mapping the source variables to the harmonized vocabulary, and (5) final inspection of the data and submission to a public repository. Each step is discussed in greater detail below under its respective subheading. The section “Step 1: Identify Data Sources” also describes the initial collection and processing procedures for the upper and lower limb accelerometry data for each study.

**Figure 1.**
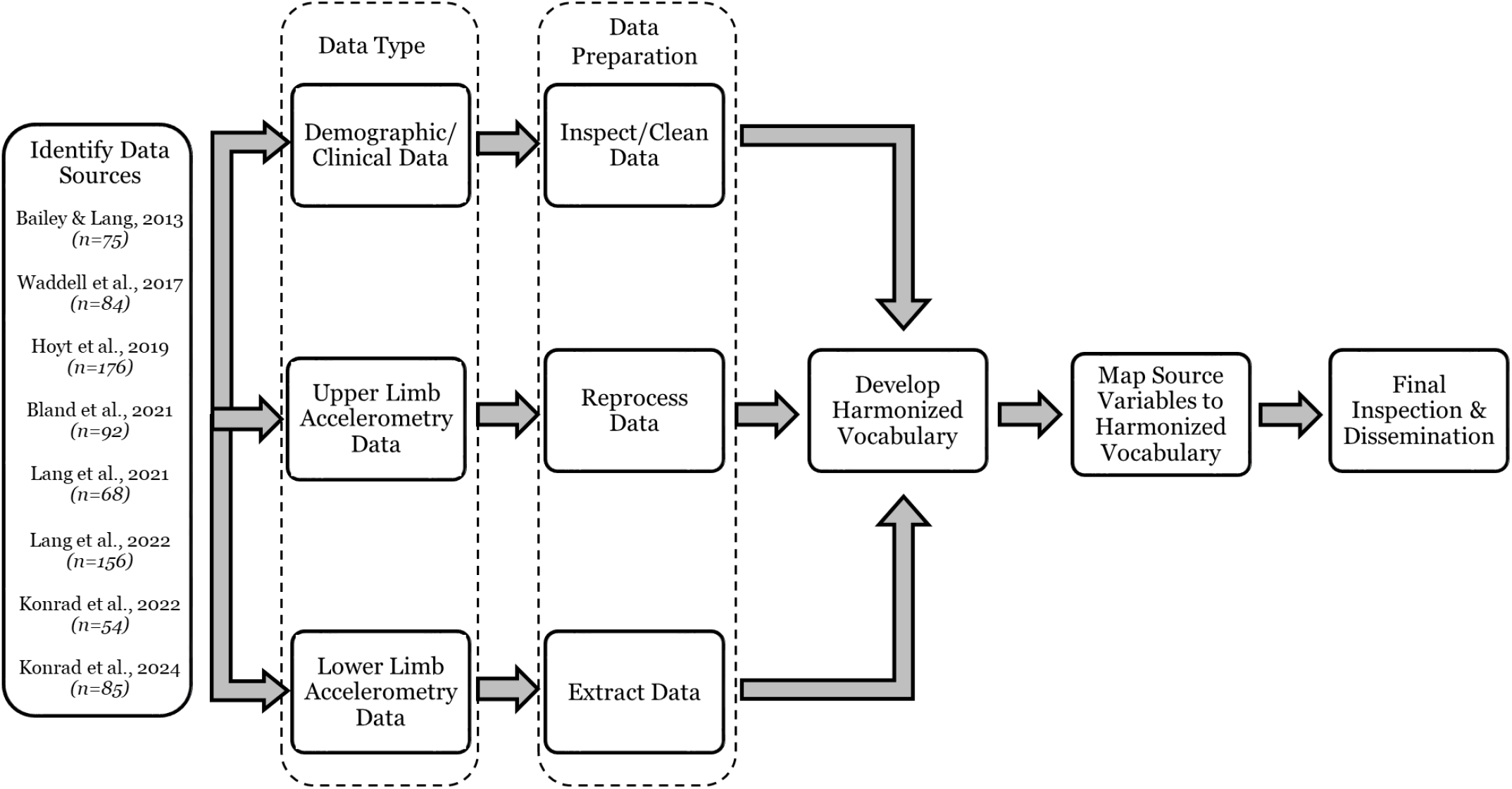
Schematic displaying how the data were harmonized. The first step involved identifying the data sources to be harmonized. The second step was to prepare the data. This step differed based on the type of data (demographic/clinical, upper limb accelerometry, lower limb accelerometry). The next steps involved developing a harmonized vocabulary and mapping the source variables to the harmonized vocabulary. The final step included inspecting the harmonized data and depositing it on a public repository.

### Step 1: Identify Data Sources

The first step was to determine which data sources to include in the harmonized dataset (Figure 1, “Identify Data Sources”). The overarching goal of the project was to harmonize accelerometry data collected by Washington University investigators and collaborators. We envisioned that users of the data would be primarily interested in answering scientific questions related to the use of accelerometry data and/or wearable sensors. Thus, the data sources that were harmonized all had to meet the following criteria: (1) include accelerometry data collected from the upper and/or lower limbs, (2) completed data collection, and (3) permit de-identified data to be shared. Eight data sources met this criterion and are listed in Figure 1 (“Identify Data Sources”). All studies were approved by Washington University Human Research Protection Office. All participants provided written informed consent. Table 1 provides the number of participants and eligibility criteria for each study as well as a brief description of the accelerometry data collection procedures.

**Table 1.**
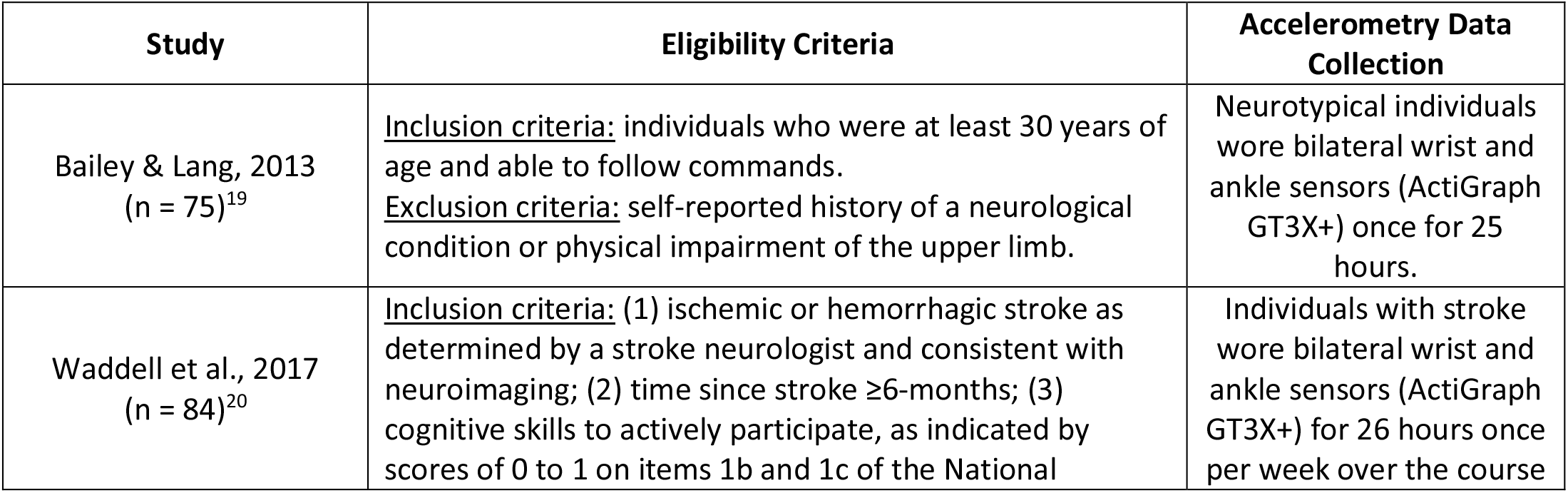
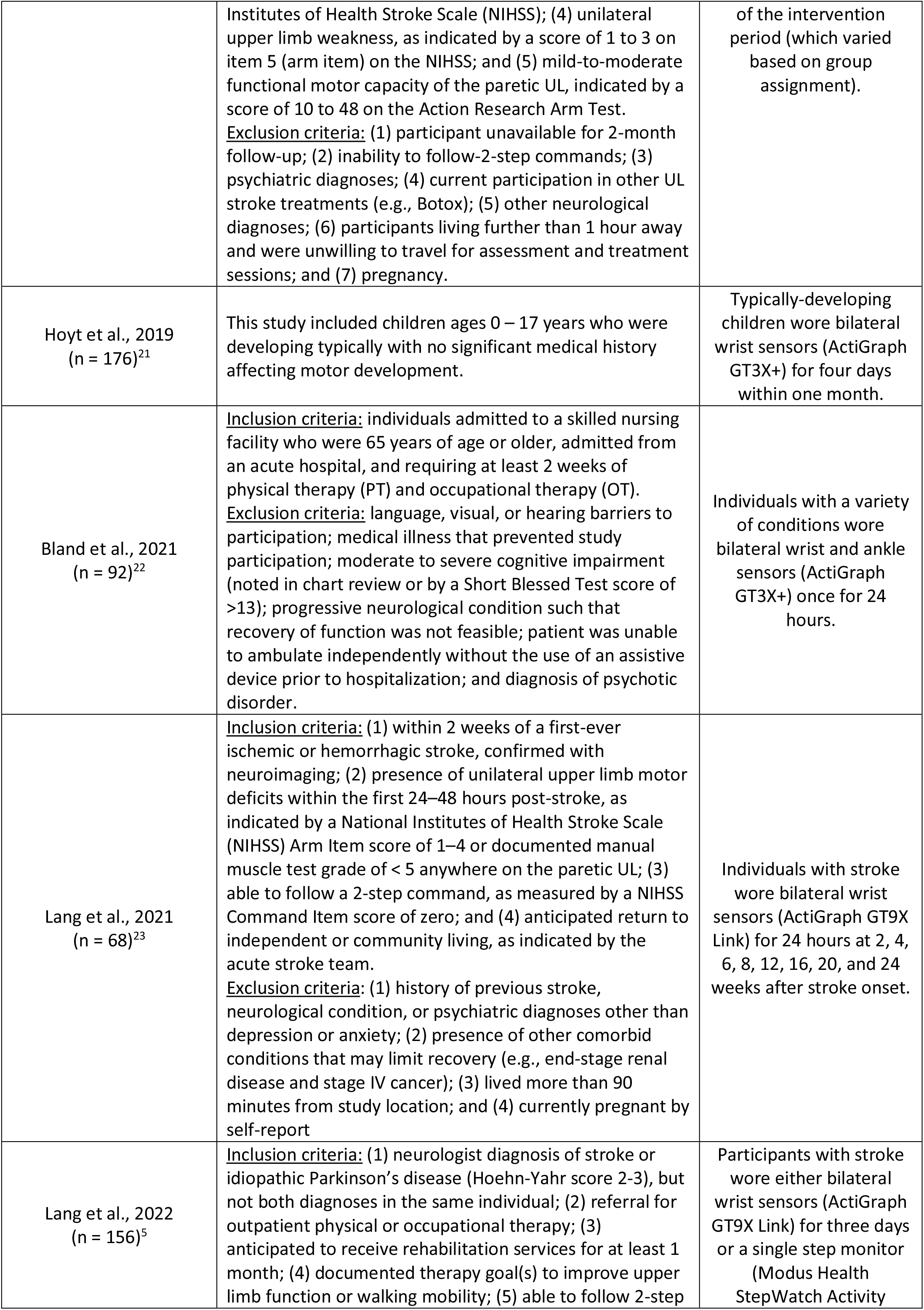
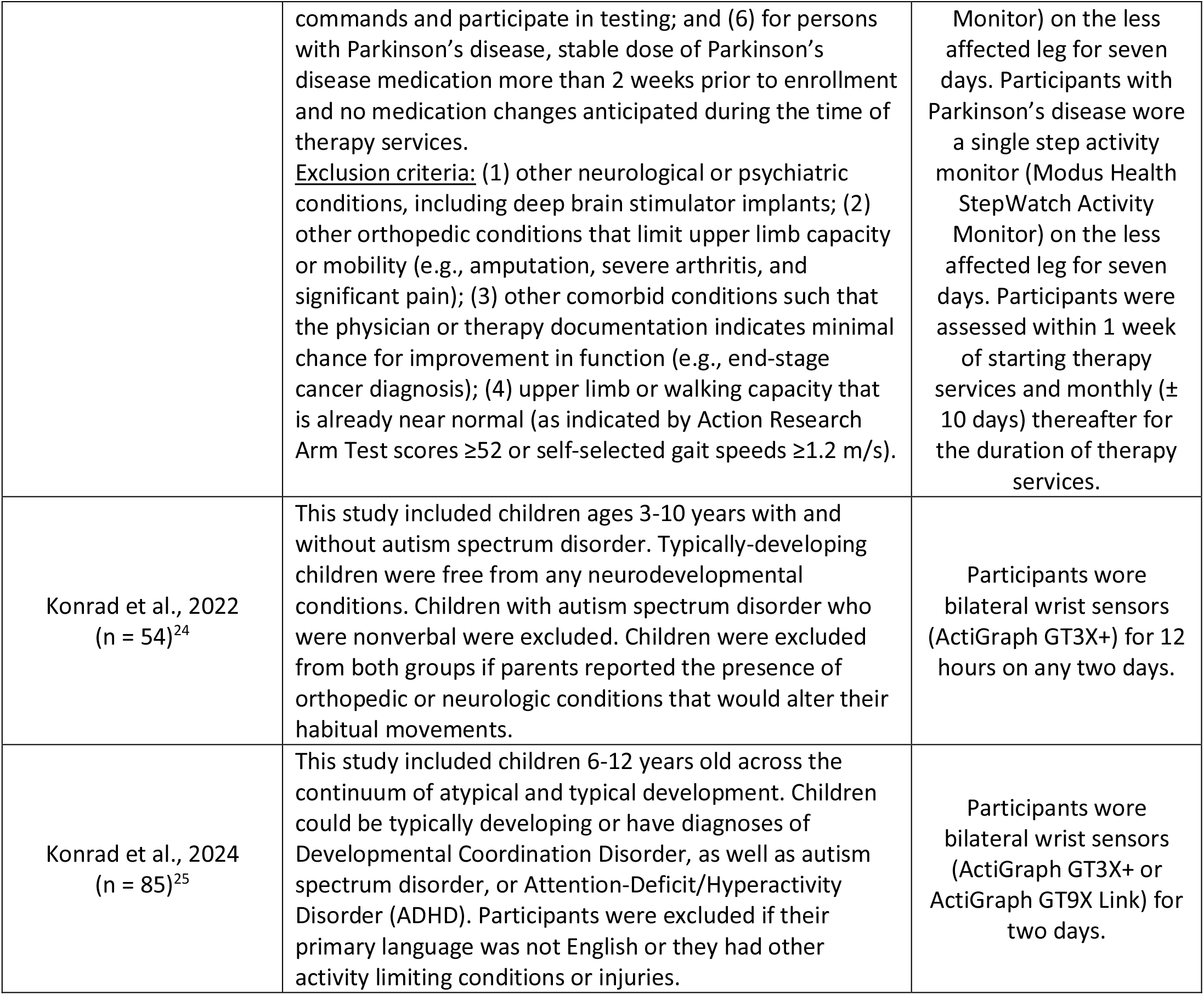
Summary of the Eight Studies Included in the Harmonized Dataset.

| Study | Eligibility Criteria | Accelerometry Data Collection |
| --- | --- | --- |
| Bailey & Lang, 2013<br>(n = 75)<sup>19</sup> | <u>Inclusion criteria:</u> individuals who were at least 30 years of age and able to follow commands.<br><u>Exclusion criteria:</u> self-reported history of a neurological condition or physical impairment of the upper limb. | Neurotypical individuals wore bilateral wrist and ankle sensors (ActiGraph GT3X+) once for 25 hours. |
| Waddell et al., 2017<br>(n = 84)<sup>20</sup> | <u>Inclusion criteria:</u> (1) ischemic or hemorrhagic stroke as determined by a stroke neurologist and consistent with neuroimaging; (2) time since stroke ≥6-months; (3) cognitive skills to actively participate, as indicated by scores of 0 to 1 on items 1b and 1c of the National | Individuals with stroke wore bilateral wrist and ankle sensors (ActiGraph GT3X+) for 26 hours once per week over the course |
|  | Institutes of Health Stroke Scale (NIHSS); (4) unilateral upper limb weakness, as indicated by a score of 1 to 3 on item 5 (arm item) on the NIHSS; and (5) mild-to-moderate functional motor capacity of the paretic UL, indicated by a score of 10 to 48 on the Action Research Arm Test.<br><u>Exclusion criteria:</u> (1) participant unavailable for 2-month follow-up; (2) inability to follow 2-step commands; (3) psychiatric diagnoses; (4) current participation in other UL stroke treatments (e.g., Botox); (5) other neurological diagnoses; (6) participants living further than 1 hour away and were unwilling to travel for assessment and treatment sessions; and (7) pregnancy. | of the intervention period (which varied based on group assignment). |
| Hoyt et al., 2019<br>(n = 176)<sup>21</sup> | This study included children ages 0 – 17 years who were developing typically with no significant medical history affecting motor development. | Typically-developing children wore bilateral wrist sensors (ActiGraph GT3X+) for four days within one month. |
| Bland et al., 2021<br>(n = 92)<sup>22</sup> | <u>Inclusion criteria:</u> individuals admitted to a skilled nursing facility who were 65 years of age or older, admitted from an acute hospital, and requiring at least 2 weeks of physical therapy (PT) and occupational therapy (OT).<br><u>Exclusion criteria:</u> language, visual, or hearing barriers to participation; medical illness that prevented study participation; moderate to severe cognitive impairment (noted in chart review or by a Short Blessed Test score of >13); progressive neurological condition such that recovery of function was not feasible; patient was unable to ambulate independently without the use of an assistive device prior to hospitalization; and diagnosis of psychotic disorder. | Individuals with a variety of conditions wore bilateral wrist and ankle sensors (ActiGraph GT3X+) once for 24 hours. |
| Lang et al., 2021<br>(n = 68)<sup>23</sup> | <u>Inclusion criteria:</u> (1) within 2 weeks of a first-ever ischemic or hemorrhagic stroke, confirmed with neuroimaging; (2) presence of unilateral upper limb motor deficits within the first 24–48 hours post-stroke, as indicated by a National Institutes of Health Stroke Scale (NIHSS) Arm Item score of 1–4 or documented manual muscle test grade of < 5 anywhere on the paretic UL; (3) able to follow a 2-step command, as measured by a NIHSS Command Item score of zero; and (4) anticipated return to independent or community living, as indicated by the acute stroke team.<br><u>Exclusion criteria:</u> (1) history of previous stroke, neurological condition, or psychiatric diagnoses other than depression or anxiety; (2) presence of other comorbid conditions that may limit recovery (e.g., end-stage renal disease and stage IV cancer); (3) lived more than 90 minutes from study location; and (4) currently pregnant by self-report | Individuals with stroke wore bilateral wrist sensors (ActiGraph GT9X Link) for 24 hours at 2, 4, 6, 8, 12, 16, 20, and 24 weeks after stroke onset. |
| Lang et al., 2022<br>(n = 156)<sup>5</sup> | <u>Inclusion criteria:</u> (1) neurologist diagnosis of stroke or idiopathic Parkinson's disease (Hoehn-Yahr score 2-3), but not both diagnoses in the same individual; (2) referral for outpatient physical or occupational therapy; (3) anticipated to receive rehabilitation services for at least 1 month; (4) documented therapy goal(s) to improve upper limb function or walking mobility; (5) able to follow 2-step | Participants with stroke wore either bilateral wrist sensors (ActiGraph GT9X Link) for three days or a single step monitor (Modus Health StepWatch Activity |
|  | commands and participate in testing; and (6) for persons with Parkinson's disease, stable dose of Parkinson's disease medication more than 2 weeks prior to enrollment and no medication changes anticipated during the time of therapy services.<br><u>Exclusion criteria:</u> (1) other neurological or psychiatric conditions, including deep brain stimulator implants; (2) other orthopedic conditions that limit upper limb capacity or mobility (e.g., amputation, severe arthritis, and significant pain); (3) other comorbid conditions such that the physician or therapy documentation indicates minimal chance for improvement in function (e.g., end-stage cancer diagnosis); (4) upper limb or walking capacity that is already near normal (as indicated by Action Research Arm Test scores $\geq 52$ or self-selected gait speeds $\geq 1.2$ m/s). | Monitor) on the less affected leg for seven days. Participants with Parkinson's disease wore a single step activity monitor (Modus Health StepWatch Activity Monitor) on the less affected leg for seven days. Participants were assessed within 1 week of starting therapy services and monthly ($\pm 10$ days) thereafter for the duration of therapy services. |
| Konrad et al., 2022<br>(n = 54)<sup>24</sup> | This study included children ages 3-10 years with and without autism spectrum disorder. Typically-developing children were free from any neurodevelopmental conditions. Children with autism spectrum disorder who were nonverbal were excluded. Children were excluded from both groups if parents reported the presence of orthopedic or neurologic conditions that would alter their habitual movements. | Participants wore bilateral wrist sensors (ActiGraph GT3X+) for 12 hours on any two days. |
| Konrad et al., 2024<br>(n = 85)<sup>25</sup> | This study included children 6-12 years old across the continuum of atypical and typical development. Children could be typically developing or have diagnoses of Developmental Coordination Disorder, as well as autism spectrum disorder, or Attention-Deficit/Hyperactivity Disorder (ADHD). Participants were excluded if their primary language was not English or they had other activity limiting conditions or injuries. | Participants wore bilateral wrist sensors (ActiGraph GT3X+ or ActiGraph GT9X Link) for two days. |

#### Accelerometry Data Collection Methods

This section describes how upper and lower limb accelerometry data were originally collected for each study.

### Upper Limb Accelerometry

All or a subset of participants in each of the eight studies were instructed to wear bilateral wrist accelerometers to measure upper limb movement. Wear times varied based on the study protocol (Table 1, “Accelerometry Data Collection”). However, the same overall methodology was applied. This methodology is described in detail in Lang et al, *J Vis Exp* 2017^14^ and summarized in this section.

Participants wore two ActiGraph GT3X+ or ActiGraph GT9X Link triaxial accelerometers (ActiGraph Corp, Pensacola, Florida), one on each wrist (Figure 2A). The same device model was used on both wrists. In the case of longitudinal studies, the same device model was used at all time points. A member of the research team met with the participant to instruct them on how to wear the accelerometers according to the study protocol. Participants were instructed to go about their usual activities and to remove the sensors during swimming activities. Participated were asked to complete a wearing log to indicate times in which the sensors were worn and removed (if necessary). Participants were provided the option to return the accelerometers either in person or a pre-paid mail envelope. The devices recorded accelerations along three axes and were programmed to record data at a frequency of 30 Hz. Once the accelerometers were returned to the lab, data were downloaded and visually inspected using ActiLife software (version 6 ActiGraph Corp, Pensacola, Florida). Data were then exported from ActiLife for further processing. The set of variables computed from the raw accelerometry data varied slightly based on the study protocol. Data were stored securely and in separate locations on Washington University’s secure Box server.

**Figure 2.**
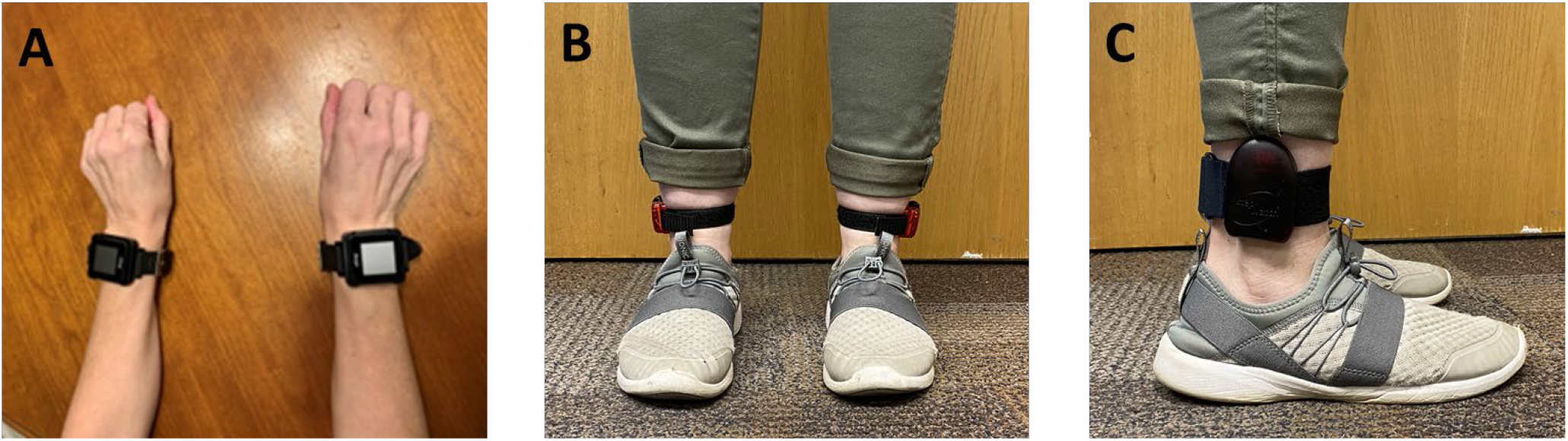
Wearable sensor placements for included studies. **A)** Participants in all studies wore ActiGraph GT3X+ or ActiGraph GT9X Link sensors on each wrist (picture shows Link sensors). **B)** Participants in three studies (Bailey et al., 2013, Waddell et al., 2017, and Bland et al., 2021) also wore bilateral ankle sensors (ActiGraph GT3X+). **C)** In one study (Lang et al., 2022), individuals with Parkinson’s disease and a subset of individuals with stroke wore a Modus StepWatch Activity Monitor (Modus Health LLC) on their less affected ankle.

#### Lower Limb Accelerometry

Four studies deployed wearable sensors worn at one or both ankle(s) to measure step counts.^5,19,20,22^ In Lang et al., 2022^5^, individuals with Parkinson’s disease and a subset of individuals with stroke were instructed to wear the Modus StepWatch Activity Monitor (Modus Health LLC) on their less affected ankle for seven days (Figure 2C).^26-28^ The device was calibrated for each participant according to the manufacturer’s instructions. The device utilizes a proprietary algorithm to measure step counts, which were extracted from the Modus StepWatch software and used to compute the average number of steps taken based on the number of recording days.

In three studies^19,20,22^, participants wore two ActiGraph GT3X+ accelerometers (ActiGraph Corp, Pensacola, Florida), one on each ankle, with wear times dictated based on the study protocol (Figure 2B). Once the accelerometers were returned to the lab, the data were processed in ActiLife software (version 6, ActiGraph Corp, Pensacola, Florida) using its proprietary algorithms to compute step counts.^29^

### Step 2: Data Preparation

Three types of data were harmonized: demographic/clinical, upper limb accelerometry, and lower limb accelerometry. These three types of data underwent different processes to prepare them for harmonization. All data sources were retrieved from Washington University’s secure Box and/or REDCap (Research Electronic Data Capture) platforms.^30,31^ Demographic/clinical data from each data source had previously been inspected and cleaned as part of its respective study protocol prior to analysis and dissemination. Here, these data underwent a second layer of inspection to verify that all values appeared reasonable and all relevant demographic/clinical variables were present (e.g., age, sex, race, and ethnicity). This was done one study at a time using both manual procedures as well as the software program, R (R Core Team, 2013, version 4.2.1). Demographic/clinical data across data sources were then merged together in Step 4 below.

Upper limb accelerometry data were reprocessed using custom-written R code (R Core Team, 2013, version 4.2.1). Reprocessing was done to ensure that all data were subject to the same processing pipeline and that the same set of variables were computed for all data sources. This process entailed extracting two data files from each wrist sensor from ActiLife software (version 6.11.9, ActiGraph Corp, Pensacola, Florida): a raw 30 Hz file (in gravitational units) and a down-sampled 1 Hz data file (in ActiGraph activity counts). The 30 Hz data were band-pass filtered from 0.2-12 Hz to remove acceleration components incompatible with human activity. Data in the 1 Hz file were first filtered using ActiGraph’s proprietary filtering algorithm, which uses a maximum gain of 0.759 Hz and goes down to -6db at 0.212 Hz at 2.148 Hz and then down-sampled from 30 Hz to 1-second epochs for each axis by summing the 30 samples within each second.^32^ Both the 1 Hz and 30 Hz files were trimmed to only include times in which the participant wore the sensors outside the clinic/lab (i.e., in daily life), which varied across studies. The exception was Bland et al., 2021^22^ in which participants were residing in a skilled nursing facility. Accelerations in each axis were combined into a single vector magnitude using the formula 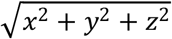. A vector magnitude threshold of ≥ 2 activity counts was used to determine if the upper limb was active for each 1-second epoch.^33,34^ For each data source, 26 variables that reflect movement of the upper limbs in daily life were computed, most from the 1 Hz data with some variables computed from the 30 Hz data (Table 2). We intentionally computed a wide range of variables for two main reasons. The first is that it is currently unknown which sensor-derived upper limb variable(s) or constructs of movement are most important to measure, and the second is that the relevance of specific variables will likely vary across research questions, clinical populations, and scientific fields.^6,21,24,35,36^ Computing a wide range of variables would therefore maximize the usefulness of this harmonized dataset for future research endeavors. The R code used to process the upper limb accelerometry variables is available on https://github.com/keithlohse/HarmonizedAccelData and archived on Zenodo.^18^ Data were reprocessed one study at a time and then merged together in Step 4 below.

**Table 2.** Upper Limb Accelerometry Variables.

| Variable Name | Variable Description | Data Source |
| --- | --- | --- |
| recording_time | The number of minutes of sensor recording time each day | 1 Hz |
| total_mvt_time<sup>37</sup> | Time (in hours) that either the left, right, or both upper limbs are moving | 1 Hz |
| l_time<sup>34</sup> | Time (in hours) that the left upper limb is moving | 1 Hz |
| l_only_time<sup>19</sup> | Time (in hours) that the left upper limb is moving, while the right upper limb is still. | 1 Hz |
| r_time<sup>34</sup> | Time (in hours) that the right upper limb is moving | 1 Hz |
| r_only_time<sup>19</sup> | Time (in hours) that the right upper limb is moving, while the left upper limb is still | 1 Hz |
| simultaneous_time<sup>37</sup> | Time (in hours) that both upper limbs are moving | 1 Hz |
| l_magnitude<sup>38</sup> | Median of the accelerations of the left upper limb, in activity counts | 1 Hz |
| r_magnitude<sup>38</sup> | Median of the accelerations of the right upper limb, in activity counts | 1 Hz |
| bilateral_magnitude<sup>38</sup> | Magnitude of accelerations of movement summed across both upper limbs, in activity counts | 1 Hz |
| l_magnitude_sd<sup>38</sup> | Standard deviation of the magnitude of accelerations across the left upper limb, in activity counts | 1 Hz |
| r_magnitude_sd<sup>38</sup> | Standard deviation of the magnitude of accelerations across the right upper limb, in activity counts | 1 Hz |
| l_peak_magnitude<sup>38</sup> | The highest magnitude of accelerations of the left upper limb, in activity counts | 1 Hz |
| r_peak_magnitude<sup>38</sup> | The highest magnitude of accelerations of the right upper limb, in activity counts | 1 Hz |
| use_ratio<sup>33</sup> | Ratio of hours of non-dominant/affected upper limb movement, relative to hours of dominant/unaffected upper limb movement | 1 Hz |
| variation_ratio<sup>38</sup> | Ratio of the standard deviation of the non-dominant/affected upper limb relative to the dominant/unaffected upper limb | 1 Hz |
| simple_magnitude_ratio<sup>38</sup> | Ratio of the magnitude of accelerations of the non-dominant/affected upper limb relative to the dominant/unaffected upper limb | 1 Hz |
| l_entropy<sup>39</sup> | A measure of the time series variability from the accelerations of the left upper limb during the hour of maximum activity. Higher values indicate a more random signal. | 1 Hz |
| r_entropy<sup>39</sup> | A measure of the time series variability from the accelerations of the right upper limb during the hour of maximum activity. Higher values indicate a more random signal. | 1 Hz |
| l_jerk_ave<sup>40</sup> | The average jerk of the left upper limb, in gravitational units/second. Higher values indicate less smooth movement. | 30 Hz |
| r_jerk_ave<sup>40</sup> | The average jerk of the right upper limb, in gravitational units/second. Higher values indicate less smooth movement. | 30 Hz |
| jerk_aym<sup>40</sup> | The ratio of the average jerk magnitude between the non-dominant/affected upper limb and the dominant/unaffected upper limb. | 30 Hz |
| l_mean_freq<sup>41</sup> | The weighted mean of the component frequencies from the acceleration time series from the left upper limb. | 30 Hz |
| r_mean_freq<sup>41</sup> | The weighted mean of the component frequencies from the acceleration time series from the right upper limb. | 30 Hz |
| l_sd_freq<sup>41</sup> | The weighted standard deviation of the component frequencies from the acceleration time series from the left upper limb. | 30 Hz |
| r_sd_freq<sup>41</sup> | The weighted standard deviation of the component frequencies from the acceleration time series from the right upper limb. | 30 Hz |

In three studies^19,20,22^, raw data from the lower limb accelerometers were processed in ActiLife software (version 6.11.9, ActiGraph Corp, Pensacola, Florida) using its proprietary step count algorithm.^29^ Ten-second epoch.agd files (ActiLife’s native file format) were uploaded to ActiLife software and step counts for each calendar day of data collection were extracted. In Lang et al., 2022^5^, the Modus StepWatch Activity Monitor (Modus Health LLC) and its proprietary software were used to measure and compute step counts. ^26-28^ The primary variable computed from the lower limb accelerometry data was average steps per day, which was calculated by summing the number of steps taken over the recording period and dividing this sum by the number of recording days. Table 3 describes the relevant variables associated with the lower limb accelerometry data. Step data were extracted one study at a time and then merged together in Step 4 below.

**Table 3.** Lower Limb Accelerometry Variables

| Variable | Description |
| --- | --- |
| SensorLocation | Indicator for limb placement of the sensor |
| AverageStepsPerDay<sup>42</sup> | This variable is computed by summing the total number of steps taken over the recording period and dividing this sum by the number of recording days (RecordingDays) |
| RecordingDays<sup>16</sup> | The number of recording days in which the participant wore the sensor; used in the calculation of AverageStepsPerDay |
| Sensor | The model of sensor used to collect the data |

### Step 3: Develop a Harmonized Vocabulary

There were similarities and differences in how variables common across data sources were named and coded. The purpose of this step was to establish a consistent variable naming and coding convention across data sources. Where possible, we utilized common data elements found in the National Institutes for Health Common Data Element Repository (https://cde.nlm.nih.gov/home). When a common data element was not available for a specific variable, it was named according to the convention used in the most recent study protocol. For example, the variable designating the participant’s sex differed in name and coding scheme across data sources. We utilized the National Institute of Neurological Disorders and Stroke Common Data Element for this variable (BirthSexAssignTyp, C58676) to ensure this variable would have the same name and coding scheme across data sources in which a value of “1” would indicate “Male” and a value of “2” would indicate “Female” for all data sources.

### Step 4: Map Source Variables to the Harmonized Vocabulary

The purpose of this step was to convert variable names and codes of the source data to the harmonized convention. The data dictionary for each study was extracted and merged into a single Excel file with a separate tab for each study. For each study and variable, the source variable name and code were mapped to the harmonized variable name and code. This process was done one study at a time, such that the end result was an Excel file with eight tabs that listed the source variable name and code and the harmonized variable name and code for each variable in the study. An example of this process is shown in Table 4.

**Table 4.** Example of the Mapping Process

| Data Source | Source Variable Name | Source Variable Description | Source Variable Code | Common Data Element ID | Harmonized Variable Name | Harmonized Variable Code |
| --- | --- | --- | --- | --- | --- | --- |
| Lang et al., 2021<sup>23</sup> | Sex | The participant’s self-reported sex | 1 Female<br>2 Male | C58676 | BirthSexAssignTyp | 1 Male<br>2 Female |

Once this process was complete, the Excel file was used as a reference to implement the variable mapping in R (R Core Team, 2013, version 4.2.1). This was done individually for each study and data type (demographic/clinical, upper limb accelerometry, lower limb accelerometry). For each study, the original data file was loaded into R and the source variable names and codes were converted to the harmonized variable names and codes. Once this conversion was complete, the data were saved and exported as a new file to preserve the original data file for quality inspection in Step 5 below. After completing this process for all data sources and types, the data sources were merged together to create a master harmonized data file that had consistent variable names and coding schemes across studies. The end result of this process was three separate harmonized csv files, one for each type of data (demographic/clinical, upper limb accelerometry, lower limb accelerometry). Each data file is organized in long format as studies varied in the number of days and time points in which accelerometry data were collected.

### Step 5: Final Inspection and Dissemination

All three data files underwent a final layer of inspection prior to submitting the data to a public repository. This inspection process included uploading each data file to R (R Core Team, 2013, version 4.2.1) and computing descriptive statistics, such as variable means, standard deviations, minimum and maximum values, to ensure all values appeared reasonable. Plots of variables were generated to visually inspect the data. Table 5 provides descriptive statistics of the harmonized sample.

**Table 5.** Demographic and Clinical Characteristics of Harmonized Sample

|  |  |
| --- | --- |
| **Total N** | 790 |
| **Total Number of Accelerometry Recording Days** | 2885 |
| **Age (years)** | 42.7 ± 30.1 (range 0 – 90) |
| **Sex** | 53% male, 47% female |
| **Race** | 69.4% White, 24.9% Black, 1.8% Asian, 0.1% American Indian or Alaska Native, 1.1% Multi Race, 2.7% Unknown |
| **Conditions** | 46% Neurotypical, 31% Stroke, 7% Parkinson’s disease, 6% Orthopedic, 3% autism spectrum disorder, 7% other medical conditions |

After all data were inspected, the final step was to deposit the data on a public repository. The Eunice Kennedy Shriver National Institute of Child Health and Human Development (NICHD) Data and Specimen Hub (DASH) repository was chosen as most studies included in the harmonized dataset were funded by NICHD. Prior to submission to DASH, the dataset was stripped of all 18 HIPAA identifiers according to the Safe Harbor method.^43^

## Data Records

The full harmonized dataset is publicly available on the NICHD DASH repository as two separate studies: Harmonized Upper and Lower Limb Accelerometry Data_Part1 (doi: https://dash.nichd.nih.gov/study/426315)^44^ and Harmonized Upper and Lower Limb Accelerometry Data_Part2 (doi: https://dash.nichd.nih.gov/study/426433)^45^. Data were required to be submitted to NICHD DASH as two separate studies due to differences in data use limitations, in which use of data from Part1 is limited to research on neurological conditions and/or movement disorders and use of data from Part2 has no additional limitations. Harmonized Upper and Lower Limb Accelerometry Data_Part 1^44^ includes the studies Lang et al., 2021^23^, Lang et al., 2022^5^, Waddell et al., 2017^20^, Bland et al., 2021^22^, Bailey & Lang, 2013^19^, and Konrad et al., 2024^25^. Harmonized Upper and Lower Limb Accelerometry Data_Part2^45^ includes the remaining two studies Hoyt et al., 2019^21^ and Konrad et al., 2022^24^. The Part1 and Part2 studies are linked in DASH, making it easy for users interested in the full harmonized dataset to locate and view both studies. Data are organized as separate csv files based on the type of data: demographic/clinical data, upper limb accelerometry data, and lower limb accelerometry data. The data are accompanied by a codebook, copies of data collection instruments, and study protocol to maximize the usability of the data. The upper and lower limb accelerometry data files contain processed, summary variables (see Tables 2 and 3). Raw data files from several of the included studies were previously made available on the SimTK repository (https://simtk.org/projects/referentaccdata). The remainder of the raw data files will be made publicly available on DASH in the near future. Users who are interested in obtaining these raw data files may contact Catherine Lang during this interim period.

## Technical Validation

All data were inspected upon initial collection according to each study protocol. Two to three research team members inspected the data again both before and after harmonization. Summary statistics of all variables were computed, including means, standard deviations, minimum and maximum values, and frequency statistics to verify that all values appeared reasonable. Data were also visualized using histograms, scatter plots, and bar graphs to inspect distributions and check for implausible values. The original values of the clinical and demographic data from each data source were compared with the harmonized dataset to verify that the mapping process was successful. For each study, we also computed descriptive statistics for the demographic (e.g., age, sex, race, ethnicity) and clinical variables (e.g., time since onset of condition) and compared these values to those reported in the primary publication associated with each data source. For example, in Bland et al., 2021^22^, the mean and standard deviation of the age of the sample reported in Table 1 are 79.5 ± 8.3 years. The mean age and standard deviation for this study in the harmonized dataset were computed and found to be the same values. This provided evidence that the harmonization process was effective at merging the data while preserving the original values. The reprocessed upper limb accelerometry data were inspected by 2-3 research team members and compared to original values, when available. These values were also the same or very similar. All research team members viewed the data and provided feedback prior to submitting it to the NICHD DASH repository.

## Data Availability

Harmonized data are available on the Eunice Kennedy Shriver National Institute of Child Health and Human Development (NICHD) Data and Specimen Hub (DASH) repository.

https://dash.nichd.nih.gov/study/426315

https://dash.nichd.nih.gov/study/426433

## Usage Notes

The harmonized data can be accessed under the NICHD DASH User Agreement, which requires users to agree to use the data for scientific research and not for commercial purposes. Use of data from Part1^44^ is also limited to research on neurological conditions and/or movement disorders. Users of these data should properly acknowledge the contributions of Dr. Catherine Lang, her team, and Washington University in all written, visual, or oral public disclosures concerning the recipient’s use of the data by citing this article and the data used (Part1^44^, Part2^45^, or both^44,45^).

Although data were subject to extensive quality control procedures, errors can still occur. Users of the data that encounter errors or wish to provide feedback can do so by contacting Catherine Lang at or (314) 286-1945. Any changes or updates to the data will be released under a new version in the NICHD DASH repository.

We have also developed an R Shiny app to allow others to visualize and interact with the dataset: https://langlab.shinyapps.io/harmonized_data/. This tool may be helpful for a variety of scenarios, including generating new scientific questions, determining which accelerometry variables may be most relevant for the user’s scientific question, and for helping interested persons make informed decisions about whether to request access to the full dataset on NICHD DASH. The R Shiny app development is ongoing.

Ultimately, we hope that this harmonized dataset will accelerate rehabilitation research by allowing the field to address important scientific questions using a large pre-existing dataset of wearable sensor data, improve the reproducibility and replicability of wearable sensor studies in rehabilitation, and minimize costs and duplicated scientific efforts along the way. An example of this is the first publication associated with these data that generated referent upper limb accelerometry data in typically-developing children.^46^ Investigators using wearable sensors to measure upper limb activity in children with movement impairments could use these referent data to compare their outcomes to those reported in this typically-developing cohort, saving costs and resources associated with recruiting a control group.

## Code Availability

Upper limb accelerometry data were processed using custom-written R code (version 4.2.1, R Core Team, 2013). Eight code scripts (one for each study) are available at https://github.com/keithlohse/HarmonizedAccelData and archived on Zenodo.^18^ All code scripts calculated the upper limb sensor variables the same way; however, variations in the number of recording days and in-lab versus out-of-lab time for each study protocol resulted in some differences in processing procedures across studies. Executing these code scripts requires two data files from each wrist sensor (four files in total) from ActiLife software (ActiGraph Corp, Pensacola, Florida): a raw 30 Hz file (in gravitational units) and a down-sampled 1 Hz data file (in ActiGraph activity counts). The code outputs the sensor variables listed in Table 2 for each recording day.

## Acknowledgements

The studies by *Bailey et al*., *2013, Waddell et al*., *2017, Lang et al*., *2021*, and *Lang et al*., *2022* were funded by NIH R01/R37 HD068290. *Konrad et al*., *2022* and *Konrad et al*., *2024* were supported in part by the Foundation for Physical Therapy Research Promotion of Doctoral Studies I Scholarship (JDK). The study by *Bland et al*., *2021* was funded by NIH R01 MH099011A1 and the Washington University Institute of Clinical and Translational Sciences grant UL1TR002345 from the National Center for Advancing Translational Sciences of the NIH. *Hoyt et al*., *2019* was funded by the Hope Center for Neurological Disorders. AEM received support and training to harmonize the data from the NIH NICHD/NCMRR R25HD105583 (ReproRehab) program.

## Author contributions

- AEM – conceptualization, data curation, methodology, software, validation, writing – original draft preparation, writing – reviewing and editing
- KRL – conceptualization, methodology, supervision, software, writing – reviewing and editing, funding acquisition
- MDB – writing- reviewing and editing; validation
- JDK – writing- reviewing and editing, software
- CRH – writing- reviewing and editing
- EJL – writing- reviewing and editing
- CEL – conceptualization, methodology, supervision, funding acquisition writing – reviewing and editing

